# Sex Difference in Causes and Timing of One-Year Outcomes Among Young Acute Myocardial Infarction Patients; Results from the VIRGO Study

**DOI:** 10.1101/2022.09.30.22280298

**Authors:** Mitsuaki Sawano, Yuan Lu, Cesar Caraballo, Shiwani Mahajan, Rachel Dreyer, Judith H. Lichtman, Gail D’Onofrio, Erica Spatz, Rohan Khera, Oyere Onuma, Karthik Murugiah, John A. Spertus, Harlan M. Krumholz

## Abstract

**Background:** Younger women have higher recurrent hospitalization rates and worse health status than men after their index episode of acute myocardial infarction (AMI). However, whether women have a higher risk of cardiovascular events in the year after discharge is unknown.

**Methods:** We used data from the VIRGO (Variation in Recovery: Role of Gender on Outcomes of Young AMI Patients) study, which enrolled young AMI patients aged 18 to 55 years across 103 US hospitals. Sex differences in all-cause and cause-specific hospitalizations were compared by calculating incidence rates (IR, per 1,000 person-years) and incidence rate ratios (IRRs) with 95% confidence intervals (CIs). We then performed sequential modeling to evaluate the sex difference by calculating sub-distribution hazard ratios (SHR) accounting for deaths.

**Results:** Among 2,007 women and 972 men, at least one all-cause hospitalization occurred among 905 (30.4%) participants in the year after discharge. The leading causes of hospitalization were MI-related (IR 171.8, 95% CI, 153.6-192.2 among women vs. IR 117.8, 95% CI, 97.3-142.6 among men), followed by non-cardiac (IR 145.8, 95% CI, 129.2-164.5 among women vs. IR 69.6, 95% CI, 54.5-88.9 among men) and other cardiac or stroke hospitalizations (IR 58.8, 95% CI, 48.8-70.7 among women vs. IR 53.8, 95% CI, 40.8-71.0 among men). Competing risk analysis showed that the sex difference was present for MI-related hospitalizations (SHR 1.33, 95%CI 1.04-1.70; P=0.02) and non-cardiac hospitalizations (SHR 1.51, 95%CI 1.13-2.07; P=0.01).

**Conclusions:** Young women with AMI have more adverse outcomes compared with men in the year after discharge. MI-related hospitalizations were the most common cause of hospitalizations, but non-cardiac hospitalizations showed the most significant sex disparity. Further studies to better understand the underlying mechanisms of non-cardiac hospitalizations are warranted.

## Introduction

Young women hospitalized for acute myocardial infarction have higher in-hospital mortality^1 2^ compared with similarly aged men, and this disparity may be worsening over time.^3^ Moreover, after the index episode of AMI, prior work has shown that younger women have higher recurrent hospitalization rates and worse health status than men.^4^ What is not yet established is the pattern of sex differences in adverse events that occur in the year after discharge. ^5,6^

A key question is whether women have an increased risk for certain cardiovascular events, complications of treatments, or non-related non-cardiovascular events. The pattern of these events can reveal opportunities to improve care, strengthen preventive strategies, and promote better health. Prior studies have indicated that young women with AMI have more comorbidity and lower health status than men.^7,8^ Consequently, acute events in the subsequent year could be more related to comorbidity than to cardiac events related to the AMI. Conversely, it is possible that recurrent, related cardiovascular events are the most common acute events for young women with AMI. This information has relevance to prevention efforts for these patients.

Accordingly, we used data from the prospective, multicenter VIRGO (Variation in Recovery: Role of Gender on Outcomes of Young AMI Patients) study to determine sex differences in causes and timing of one-year adverse events after AMI in people under 55 years to identify targets for interventions and mitigate sex-disparities in cardiovascular care and outcomes. The VIRGO study has recently completed full adjudication of the cause of acute events in the year after discharge and is now ideally positioned to investigate sex differences in one-year hospitalization events after an AMI.

## Methods

### Study population

The design of the VIRGO study has been described in detail elsewhere.^9^ In brief, the VIRGO study is an observational study of the presentation, treatment, and outcomes of young women and men with AMI aged 18 to 55. The study included patients with AMI in 103 geographically diverse hospitals in the United States from August 2008 to January 2012 using a strict 2:1 enrollment ratio of women to men. The study also included patients from Spain and Australia, but outcomes adjudication could only be performed in the US cohort. Institutional review board approval was obtained at each participating institution, and patients provided informed consent for their study participation, including baseline hospitalization and follow-up interviews. For the current study, 2,985 US patients (N=2,009 women, N=976 men) hospitalized for AMI were included. After excluding in-hospital deaths (N=6), this resulted in a cohort of 2,979 participants.

### Definitions of Key Variables

Race and ethnicity were self-reported by study participants, as non-Hispanic American Indian or Alaska Native, non-Hispanic Asian or Pacific Islander, Hispanic or Latino, non-Hispanic Black, non-Hispanic White, or a combination of these. AMI was defined as (1) an increase in cardiac biomarkers (troponin I or T or creatine kinase-MB) with at least 1 value >99th percentile of the upper reference limit within 24 hours of admission and (2) supporting evidence of acute myocardial ischemia, including symptoms or ECG changes. Type of MI were classified using the VIRGO taxonomy.^10^ Health status included the Short Form-12 score (Physical and mental component scores), Euro-QoL Scale (EQ-5D) score, Seattle Angina Questionnaire (overall and individual domain scores), Patient Health Questionnaire (PHQ)-9, ENRICHD Social Support Instrument (ESSI), Perceived stress scale (PSS) and were evaluated at three independent time-points: baseline, 1-month post discharge, and 12 months after discharge. We collected information on procedures performed during initial hospitalization and after discharge that included performance of coronary catheterization, angioplasty, stent placement, coronary artery bypass graft surgery, pacemaker insertion, or intracardiac defibrillator placement.

### Study Data Adjudication and Outcome Definitions

As described previously, mortality events were ascertained through interviews with family members and verified with death certificates, hospital records, or obituaries. All hospitalizations records were collected through a two-stage process.^8^ First, the research coordinator at the local site reviewed the records within their hospital network to identify hospitalization records. In addition, the study participants were asked to self-report any hospitalizations during their 1-year post-AMI interviews. Second, the Yale Coordinating Center reconciled the hospital records with participants’ self-reported events to ensure that no hospitalizations were missed. The Yale Coordinating Center requested the missing records from hospitals outside of the site networks when necessary. Further, event adjudications were completed by 5 physicians and an advanced practice registered nurse. The first 253 hospitalizations were double adjudicated, and subsequent hospitalizations underwent single adjudication. Discrepancies between adjudicators were resolved by consensus including an additional physician when necessary.

The primary outcomes of this study were all-cause and cause-specific acute events requiring hospitalization, defined as any hospital or observation stay greater than 24 hours within 1 year of discharge. These events were categorized into three groups: MI-related hospitalization, other cardiac or stroke hospitalization, and non-cardiac hospitalization. MI-related events were defined as a composite of hospitalizations due to recurrent AMI and stable/unstable angina. Other cardiac or stroke events were defined as a composite of heart failure, other cardiac causes (pericarditis, valvular disease, arrhythmias) and stroke. Non-cardiac hospitalizations included any cause of hospitalization or visit that was not classified as cardiac or hospitalizations due to diabetic complications.

### Statistical analysis

We compared baseline variables between women and men using Chi-square or Fisher’s exact test for categorical variables and student’s t-tests for continuous variables. Categorical variables are presented as number (%) and continuous variables are presented as mean (standard deviation [SD]) or median (interquartile range). Incidence rates (IR) and incidence rate ratios (IRRs) with 95% confidence intervals (CIs) were calculated and compared between sexes. IR was calculated per 1,000 person-years and IRR was calculated with men as the reference, Kaplan–Meier survival probability estimates, and Log-rank tests were calculated from the discharge date to 365 days for 1-year outcomes. Variables with missing values were imputed using the multiple imputation method with 10 imputations.^11^ The actual missing values ranged from 0.03% (history of depression) to 10.1% (type of AMI).

We aimed to quantify the sex differences in 1-year outcomes, and then explore the factors contributing to such sex differences. Thus, we fit a set of Cox models by sequential adjustment for potential confounders and calculated the subdistribution hazard ratio^12^ (SHR) of sex upon 1-year outcomes (all-cause hospitalizations, MI-related, other cardiac/stroke, and non-cardiac hospitalizations). Specifically, Model 1 included only sex, Model 2 included Model 1 variable and demographic and comorbidity variables, Model 3 included Model 2 variables and psychosocial factors and health status, Model 4 included Model 3 and AMI presentation and treatment factors, Model 5 included Model 4 and pre-discharge status and prescriptions (Supplemental Table 1). Variables were selected based on clinical importance and previous studies studying long-term outcomes of patients with AMI. Presence of interaction for sex and age (below the median age 49) and sex and non-Hispanic black group were tested in Model 2. In all the models, deaths before a hospitalization were accounted for using the Fine and Gray method for competing risks.^12^ We used the Schoenfeld residuals to assess the proportional hazards assumption (Supplemental Table 2).^13^ Explained variation statistic R^2^ for survival data was calculated for each Model.^14^ All analyses were performed with Stata/IC 15.1 for Mac (StataCorp, 4905 Lakeway Dr, College Station, TX 77845). P-values were 2-sided and values <0.05 were considered statistically significant.

## Results

### Characteristics of Study Cohort

Overall, there were 2,007 women and 972 men included in the final cohort of young AMI patients (Supplemental Table 3). The mean age was 47.1±6.2, 2.5% were non-Hispanic American Indian or Alaska Native, 1.5% were non-Hispanic Asian or Pacific Islander, 7.9% were Hispanic or Latino, 17.5% were non-Hispanic Black and 70% were non-Hispanic White. As compared with men, women had a higher proportion of non-Hispanic Black individuals (women vs men, 20.6% vs 10.9%; *P*<0.001), and higher prevalence of comorbidities including obesity (55.2% vs 47.7%; *P*<0.001), chronic obstructive lung disease (14.2% vs 6.4%; *P*<0.001), congestive heart failure (5.7% vs 2.3%; *P*<0.001), prior stroke (4.1% vs 1.9%; *P*=0.001) and renal disease (12.7% vs 8.6%; *P*<0.001). Furthermore, young women had a higher proportion of low-income (47.6% vs 31.4%; *P*<0.001), history of depression at baseline (48.7% vs 24.2%; *P*<0.001) and significantly worse health status (Supplemental Table 4). Upon hospital presentation, women were less likely to present with chest pain (86.3% vs 89.2%; *P*=0.03) and experienced longer delays in hospital arrival from symptom onset (47.6% vs 38.0%; *P*<0.001) (Supplemental Table 5). Non ST-elevated myocardial infarction presentation (54.2% vs 42.1%; *P*<0.001) and myocardial infarction with nonobstructive coronary arteries (MINOCA) (12.8% vs 2.9%; *P*<0.001) were more prevalent among women. At the time of discharge, the total length of stay was longer for women, and women received lower rates of guideline-recommended medical therapies including aspirin (92.6% vs 95.0%; *P*<0.02), statins (67.5% vs 71.7%; *P*<0.001), beta-blockers (89.6% vs 94.1%; *P*<0.001), and angiotensin-converting enzyme inhibitors (ACEi)/angiotensin receptor blockers (ARBs) (61.2% vs 70.6%; *P*<0.001).

### Sex difference in 1-year events

The observed rates (95% CI) of all-cause hospitalizations within 1-year after discharge were 34.8% (95% CI, 32.7%-36.9%) and 23.0% (95% CI, 20.4%-25.8%) for women and men, respectively. The leading cause of these acute events among women were MI-related with an incidence rate of 171.8 (95% CI, 153.6-192.2) per 1,000 person-years that included AMI hospitalizations (IR=30.9 [95% CI, 24.0-39.8] per 1,000 person-years) and stable or unstable angina hospitalizations (IR=135.2 [95% CI, 119.4-153.2] per 1,000 person-years), followed by non-cardiac at 145.8 (95% CI, 129.2-164.5) per 1,000 person-years and other cardiac or stroke hospitalization at 58.8 (95% CI, 48.8-70.7) per 1,000 person-years that included heart failure hospitalization (IR=25.7 (95% CI, 19.5-33.9) per 1,000 person-years) and other cardiac (IR=26.2 (95% CI, 19.9-34.6) per 1,000 person-years) (Table 1). The leading cause of the acute events among men were MI-related with an incidence rate of 117.8 (95% CI, 97.3-142.6) per 1,000 person-years that included AMI (IR=25.3 (95% CI, 17.0-37.8) per 1,000 person-years) and stable or unstable angina (IR=89.3 (95% CI, 71.8-111.0) per 1,000 person-years), followed by non-cardiac at 69.6 (95% CI, 54.5-88.9) per 1,000 person-years and other cardiac or stroke at 53.8 (95% CI, 40.8-71.0) per 1,000 person-years that included heart failure (IR=16.8 (95% CI, 10.3-27.4) per 1,000 person-years) and other cardiac cause (IR=17.2 (95% CI, 10.5-28.1) per 1,000 person-years). When compared with men, women had significantly higher IRRs for MI-related (IRR 1.46, 95%CI 1.16-1.84; *P*<0.001) and for non-cardiac events (IRR 2.10, 95%CI 1.59-2.80; *P*<0.001), with similar rates for other cardiac or stroke hospitalizations (IRR 1.09, 95%CI 0.78-1.56; *P*<0.61). Of note, the timing of hospitalizations (Supplemental Table 6) nor invasive procedures (Supplemental Table 7) performed during the hospitalizations did not greatly differ between sex. Unadjusted Kaplan-Meier survival curves showed that all-cause hospitalizations, MI-related and non-cardiac hospitalizations peaked at the first month after discharge and gradually declined to reach a steady state after 3 months after discharge for both sexes (Figures 1& 2).

**Table 1.**
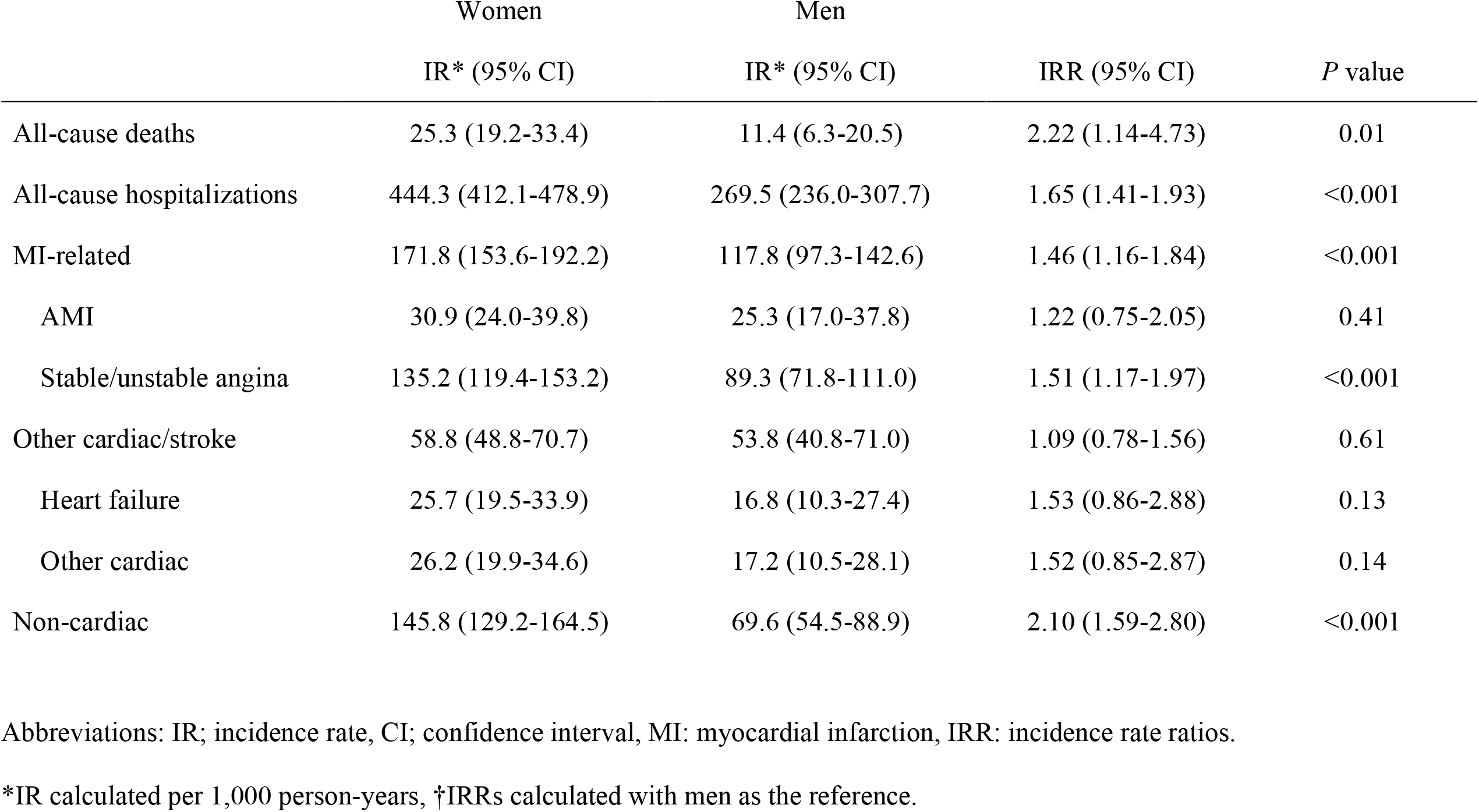
Incidence rates and incidence rate ratios for 1-year outcomes among young women and men

**Figure 1.**
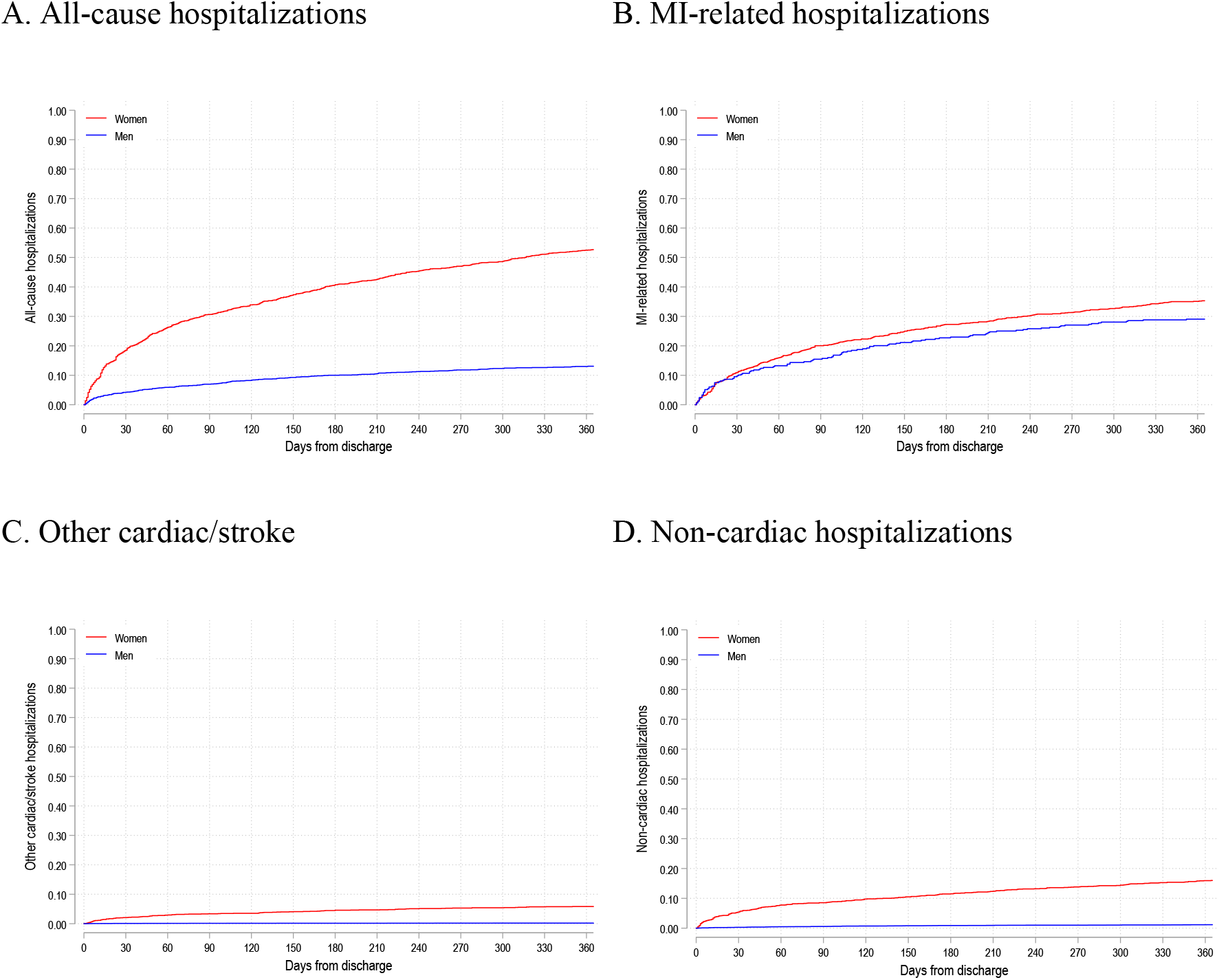
Age-adjusted Kaplan Meier curves showing 1-year all-cause hospitalizations, and cause-specific hospitalizations A, All-cause hospitalizations. B, myocardial infarction (MI) related hospitalizations. C, other cardiac/stroke hospitalizations. D, non-cardiac hospitalizations.

**Figure 2.**
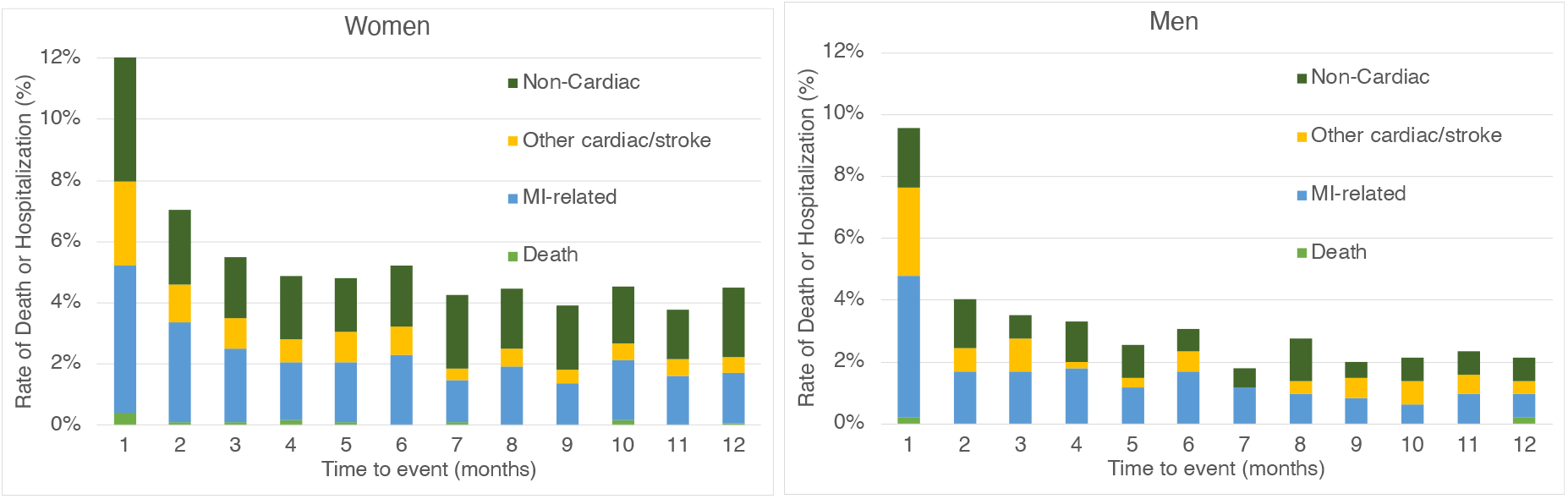
Event rate staked bar graph depicting the change in the incidence of repeat hospitalizations after AMI for different reasons over 1-month follow-up time frames among young women and men Abbreviations: MI: myocardial infarction

### Factors contributing to 1-year outcomes

Competing risk analysis adjusting for confounders demonstrated that women were consistently at significantly higher risk for 1-year all-cause, MI-related, and non-cardiac events (Figure 3). The SHRs for sex difference upon MI-related events were Model 1 (SHR 1.44, 95%CI 1.15-1.80; *P*<0.001, R^2^=0.02), Model 2 (SHR 1.43, 95%CI 1.13-1.80; *P*<0.001, R^2^= 0.06), Model 3 (SHR 1.29, 95%CI 1.01-1.64; *P*=0.04, R^2^= 0.11), Model 4 (SHR 1.33, 95%CI 1.04-1.64; *P*=0.02, R^2^=0.13) and Model 5 (SHR 1.33, 95%CI 1.04-1.70; *P*=0.02, R^2^=0.15). Of note, the addition of psychosocial factors led to an attenuation in the SHR for sex difference, while the addition of AMI presentation and predischarge treatment variables lead to minimal to no change. The SHR for sex difference non-cardiac events were Model 1 (SHR 2.06, 95%CI 1.57-2.71; *P*<0.001, R^2^=0.06), Model 2 (SHR 1.88, 95%CI 1.88-1.43; *P*<0.001, R^2^=0.11), Model 3 (SHR 1.56, 95%CI 1.17-2.07; *P*<0.001, R^2^=0.24), Model 4 (SHR 1.52, 95%CI 1.14-2.07; *P*<0.001, R^2^=0.25), and Model 5 (SHR 1.51, 95%CI 1.13-2.07; *P*=0.01, R^2^=0.29). The addition of demographics and comorbidities as well as psychosocial factors lead to attenuation in SHR for sex difference while the addition of AMI presentation and predischarge treatment variables lead to minimal to no change. No significant interactions were found for the prespecified interaction terms in any of the four outcomes.

**Figure 3.**
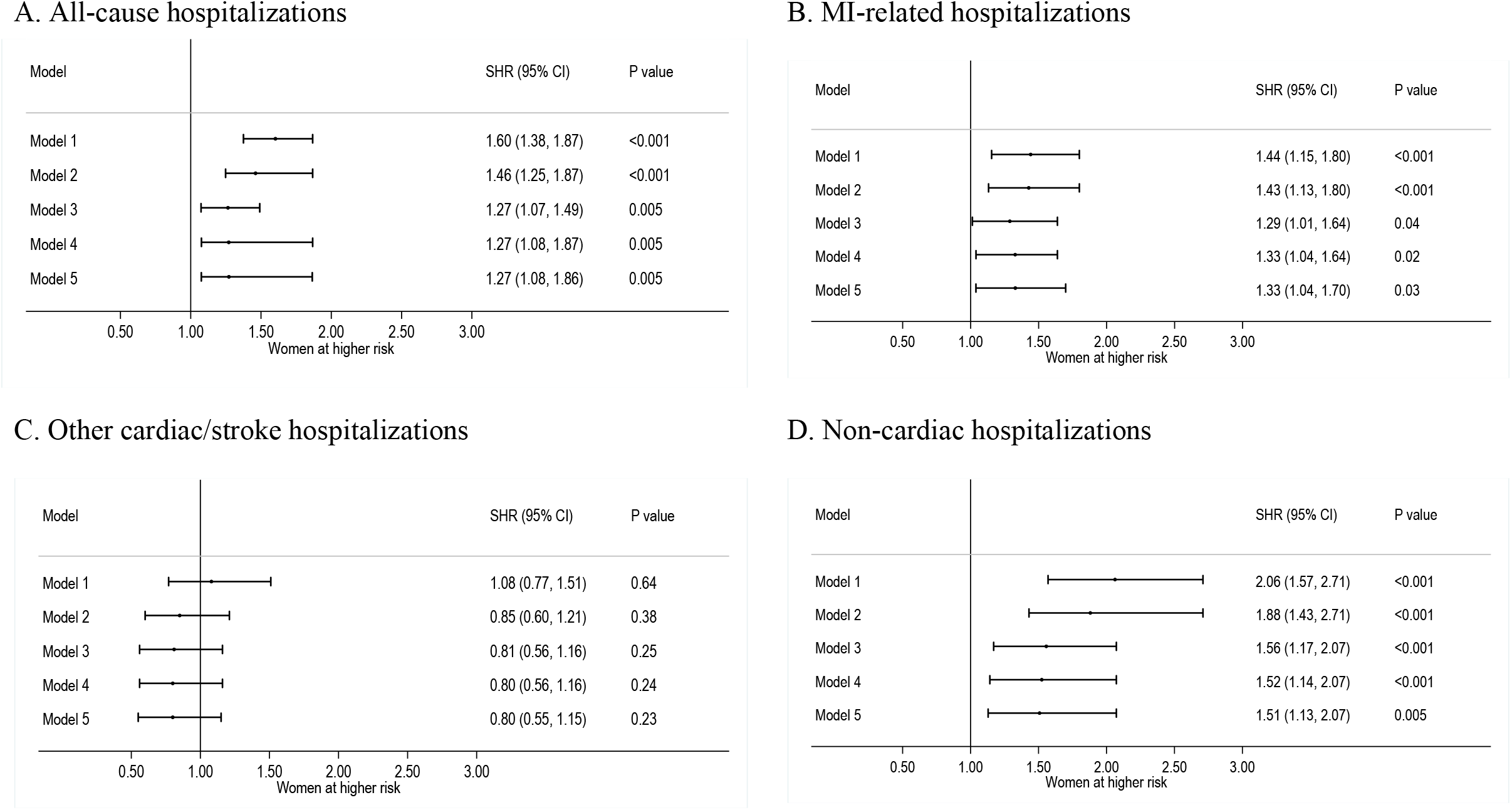
Sequential competing risk analysis for the effect of sex on 1-year outcomes A, All-cause hospitalizations. B, myocardial infarction (MI) related hospitalizations. C, other cardiac/stroke hospitalizations. D, non-cardiac hospitalizations.

## Discussion

Young women who survive an AMI have significantly more adverse events in the year after discharge compared with men. MI-related events followed by non-cardiac events were the most common cause for hospitalizations for both women and men. Further, higher risks among young women were consistently observed for all-cause, MI-related, and non-cardiac events after adjustment for potential confounders. Notably, non-cardiac events remained with greatest disparity between sex, suggesting the need for in-depth research into the causes and preventability of these non-cardiac hospitalizations to mitigate for sex differences in outcomes after an AMI.

This study extends the existing literature in several important ways. First, this is the first comprehensive assessment of sex differences in 1-year incidence of clinically important cardiovascular events including all-cause deaths and cause-specific acute events within this population of young patients with AMI. We previously reported that health status was persistently worse over the time course of 1-year among women, however, were not able to report 1-year hospitalizations since event adjudication had not been complete at the time of publication.^15^ The majority of prior studies evaluating sex difference in post-discharge AMI outcomes were conducted among older patients derived from clinical trials or administrative databases and were not primarily designed to assess young patients’ outcomes.^16, 17^ Of note, the incidence of all-cause deaths in the 1 year of discharge was low and comparable to the findings in the Partners YOUNG MI Registry^18^ but lower than those reported from the community-based surveillance conducted within the Atherosclerosis Risk in Communities (ARIC) Surveillance study.^3^ The higher rate of 1-year mortality observed in ARIC may be due to their retrospective design for collecting outcome data that may have allowed higher risk patients to be included in their study population compared to the prospectively enrolled VIRGO population.

Second, reports on specific causes of hospitalizations have been limited among patients experiencing AMI at a young age. In a study of young AMI patients with commercial health insurance collected in MarketScan, the incidence of recurrent AMI was found to be 28.4 (per 1,000 person-years) among women and 19.0 (per 1,000 person-years) among men aged between 21 to 54 years old which was lower than those observed in our registry. The difference in observed outcomes may have been due to large convenience sampling^19^ in the MarketScan dataset and due to the low to moderate sensitivity for outcome events often confronted when using administrative databases.^20^ Our ability to conduct a two-stepped event data collection process strengthened with full event adjudication is likely to have contributed to making better estimates on true post-discharge events.^9^

Third, based on rigorous adjustments for confounders, we found a consistently higher risk for MI-related and non-cardiac hospitalizations among young women. Previous VIRGO study identified sex difference and 9 other factors associated with all-cause hospitalizations but did not describe cause-specific events.^8^ Importantly, sex differences were greatest in non-cardiac hospitalizations that are consistent with a young AMI California state inpatient database showing high risk of non-cardiac hospitalizations within 30-days after discharge described as post-hospital syndrome^21,4^ In our study, this increased risk was observed prolonging over one-month and up to one year with young women at 51% greater risk compared to men. Moreover, in the sequential model, a large proportion of non-cardiac hospitalizations were explained by demographics, comorbidities, and psychosocial factors. These findings suggest the need for a greater attention to cardiac, non-cardiac, and mental health issues to mitigate this sex disparity in young women recovering from an AMI. Future research should develop and test strategies, which may include intensification of post-discharge management through in-person or virtual sessions to improve health literacy, patient motivation and address socioeconomic that may cause barriers to reach satisfactory levels of adherence to evidence-based secondary prevention strategies.^22^

### Limitations

The findings of this study should be interpreted in the context of several limitations. First, details on non-cardiac hospitalizations were not collected. Second, due to a relatively sample size, our results may not be generalizable to population groups that were underrepresented in the study. Despite this limitation it is important to note that this is the largest subset of young patients with AMI in the United States, and the composition of our cohort and prevalence of comorbidities and risk factors is consistent with other studies of younger patients with AMI.^23^ Third, only patients hospitalized with AMI who survived through acute care and provided consent were selected in this study. Selection bias towards lower risk AMI patients may have resulted in lower burden of comorbidities and lower one-year outcomes than the real-world.

## Conclusions

Young women with AMI have persistently worse outcomes compared to men immediately and 1-year after discharge. MI-related hospitalizations were the most common cause of hospitalizations, nonetheless, non-cardiac hospitalizations showed the greatest sex disparity among young patients after an AMI. Further studies to better understand the underlying mechanisms of non-cardiac hospitalizations are warranted.

## Supporting information

VIRGO Participating Sites

## Data Availability

All data produced in the present study are available upon reasonable request to the corresponding author.

## Non-standard Abbreviations and Acronyms

ACEi: angiotensin-converting enzyme inhibitor
AMI: acute myocardial infarction
ARB: angiotensin receptor blocker
ARIC: Atherosclerosis Risk in Communities
CABG: coronary artery bypass graft
CCB: Calcium channel blocker
CI: confidence interval
COPD: chronic obstructive pulmonary disease
CVD: cardiovascular disease
EQ-5D: Euro QoL 5D
ESSI: ENRICHD Social Support Inventory
ICD: intracardiac defibrillator
IR: incidence rate
IRR: incidence rate ratio
MI: myocardial infarction
MINOCA: myocardial infarction with nonobstructive coronary arteries
NSTEMI: non-ST myocardial infarction
PCI: Percutaneous coronary intervention
PHQ-9: Patient Health Questionnaire
POBA: plain old balloon angioplasty
SAQ: Seattle Angina Questionnaire
SD: standardized difference
SF-12: Short Form-12
SHR: sub-distribution hazard ratio
STEMI: ST-elevated myocardial infarction
VIRGO: Variation in Recovery: Role of Gender on Outcomes of Young AMI Patients

## Acknowledgments

We thank Dr. Yun Wang, Section of Cardiovascular Medicine, Department of Internal Medicine, Yale School of Medicine, New Haven, Connecticut, for his valuable comments on the statistical analysis

## Supplemental Materials

Supplemental Table 1-7

Supplemental Figure 1

