## Supplemental Materials for "Sex Difference in Causes and Timing of One-Year Outcomes Among Young Acute Myocardial Infarction Patients; Results from the VIRGO Study"

Supplemental Table 1. Variables included in sequential competing risk models

| Name of Model | Variables added to previous model |
|---|---|
| Model 1 | Sex |
| Model 2: Model 1 + Demographics/Comorbidities | Age, non-Hispanic black group, obesity, dyslipidemia, diabetes mellitus, COPD, previous history of MI, previous history of heart failure, previous history of stroke, renal dysfunction |
| Model 3: Model 2 + Psychosocial factors | Low level of education, low income, uninsured, history of depression, SF-12 (physical component score), SF-12 (mental component score), Seattle Angina Questionnaire-Physical limitations (SAQ), Angina frequency (SAQ) score, Treatment satisfaction (SAQ) score, Quality of life (SAQ) score, Depressive symptoms (Patient Health Questionnaire-9 score), Stress level (Perceived Stress Scale-14 score) |
| Model 4: Model 3 + AMI presentation | STEMI, Type 1 MI, presence of chest pain symptom, late presentation to the hospital after symptom onset, GRACE score |
| Model 5: Model 4 + Predischarge status and treatment | Heart failure as a complication during hospitalization, total length of stay, discharge |

| | |
|---|---|
| | medication status for aspirin, clopidogrel, statins, beta-blockers, and ACEi/ARBs |

Supplemental Table 2. Schoenfeld residuals testing the proportional-hazards assumption

| | Chi-square value | Degrees of Freedom | *P* value |
|---|---|---|---|
| All-cause hospitalizations | 44.88 | 38 | 0.20 |
| MI-related hospitalizations | 52.27 | 38 | 0.06 |
| Other cardiac/stroke hospitalizations | 39.38 | 38 | 0.41 |
| Non-cardiac hospitalizations | 40.61 | 38 | 0.36 |

Supplemental Figure 1. Log-log plot of survival after Cox regression analysis using complete data

A. All-cause hospitalizations

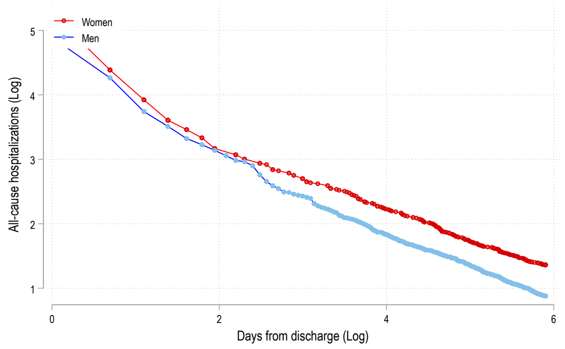


B. MI-related hospitalizations

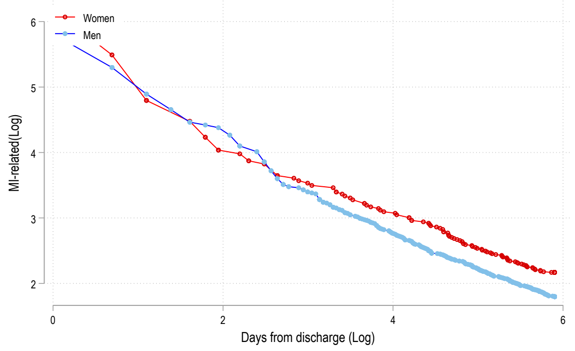


C. Other cardiac/stroke hospitalizations

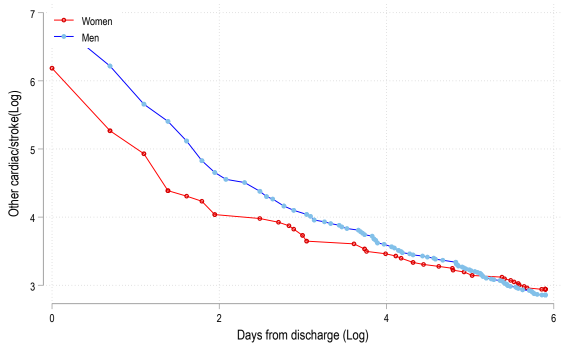


D. Non-cardiac hospitalizations

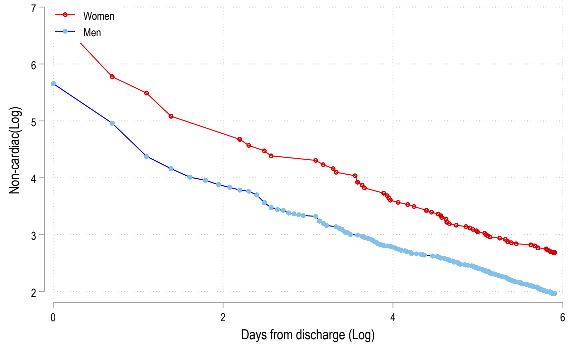


A, All-cause hospitalizations. B, myocardial infarction (MI) related hospitalizations. C, other cardiac/stroke hospitalizations. D, non-cardiac hospitalizations.

Supplemental Table 3. Sex difference in baseline demographics among young AMI patients

| | Total<br>N=2,979 | Women<br>N=2,007 | Men<br>N=972 | Standardized Difference |
|---|---|---|---|---|
| Age, mean (SD), years | 47.1 (6.2) | 47.2 (6.3) | 47.1 (5.9) | 0.02 |
| Race and Ethnicity (%) | | | | 0.29 |
| Non-Hispanic American Indian or Alaska Native | 73 (2.5) | 40 (2.0) | 33 (3.4) | |
| Non-Hispanic Asian or Pacific Islander | 44 (1.5) | 31 (1.5) | 13 (1.3) | |
| Hispanic or Latino | 235 (7.9) | 153 (7.6) | 82 (8.4) | |
| Non-Hispanic Black | 520 (17.5) | 414 (20.6) | 106 (10.9) | |
| Non-Hispanic White | 2,085 (70.0) | 1,356 (67.6) | 729 (75.0) | |
| Other | 19 (0.6) | 10 (0.5) | 9 (0.9) | |
| Obesity (%) | 1,571 (52.8) | 1,107 (55.2) | 464 (47.7) | 0.15 |
| Hypertension (%) | 1,974 (66.3) | 1,347 (67.1) | 627 (64.5) | 0.06 |
| Dyslipidemia (%) | 2,032 (68.2) | 1,331 (66.3) | 701 (72.1) | 0.27 |
| Diabetes mellitus (%) | 1,058 (35.5) | 799 (39.8) | 259 (26.6) | 0.28 |
| Smoking (%) | 891 (29.9) | 601 (29.9) | 290 (29.8) | 0.002 |
| History of COPD (%) | 346 (11.6) | 284 (14.2) | 62 (6.4) | 0.24 |
| Family History of CVD (%) | 2,004 (67.5) | 1,350 (67.5) | 654 (67.5) | 0.001 |
| Previous MI (%) | 635 (21.3) | 413 (20.6) | 222 (22.8) | 0.06 |
| Prior PCI (%) | 488 (16.4) | 306 (15.2) | 182 (18.7) | 0.00 |

| | | | | |
|---|---|---|---|---|
| Prior CABG (%) | 115 (3.9) | 77 (3.8) | 38 (3.9) | 0.17 |
| Congestive heart failure (%) | 137 (4.6) | 115 (5.7) | 22 (2.3) | 0.12 |
| Prior stroke (%) | 100 (3.4) | 82 (4.1) | 18 (1.9) | 0.03 |
| Prior PAD (%) | 74 (2.5) | 53 (2.6) | 21 (2.2) | 0.13 |
| History of renal disease (%) | 337 (11.4) | 254 (12.7) | 83 (8.6) | 0.13 |
| Physical inactivity (%) | 1,054 (35.4) | 751 (37.4) | 303 (31.2) | 0.15 |

Abbreviations: CABG, coronary artery bypass graft; COPD, chronic obstructive pulmonary disease; CVD, cardiovascular disease; MI, myocardial infarction; PCI, percutaneous coronary intervention; SD, standardized difference.

Supplemental Table 4. Sex difference in Baseline Psychosocial Factors

| | Total N=2,979 | Women N=2,007 | Men N=972 | Standardized Difference |
|---|---|---|---|---|
| Final education, High school (%) | 1,675 (56.7) | 1,130 (56.8) | 545 (56.5) | 0.004 |
| Currently working (%) | 1,828 (61.4) | 1,128 (56.2) | 700 (72.0) | 0.33 |
| Primary earner (%) | 2,214 (74.3) | 1,484 (73.9) | 730 (75.1) | 0.03 |
| Low income (%) | 1,260 (42.3) | 955 (47.6) | 305 (31.4) | 0.36 |
| Living alone (%) | 399 (13.4) | 253 (12.6) | 146 (15.0) | 0.07 |
| Has health insurance (%) | 2,294 (77.0) | 1,569 (78.2) | 725 (74.6) | 0.09 |
| History of depression (%) | 1,212 (40.7) | 977 (48.7) | 235 (24.2) | 0.51 |
| Social support (ENRICHD Social Support Instrument-7), mean (SD), points | 28.2 (5.7) | 28.1 (5.7) | 28.5 (5.7) | 0.05 |
| General health, SF-12 (physical component score), mean (SD), points | 43.0 (12.1) | 41.9 (12.2) | 45.3 (11.5) | 0.29 |
| General health, SF-12 (mental component score), mean (SD), points | 45.5 (12.4) | 44.1 (12.6) | 48.3 (11.5) | 0.34 |
| Baseline SAQ, mean (SD), points | | | | |
| Physical limitations | 80.6 (25.8) | 77.8 (27.3) | 86.3 (21.3) | 0.34 |
| Angina frequency | 83.2 (20.8) | 81.9 (21.8) | 85.7 (18.3) | 0.18 |
| Treatment satisfaction | 91.8 (13.0) | 91.1 (14.0) | 93.1 (10.7) | 0.16 |
| Quality of life | 57.4 (24.9) | 55.4 (25.7) | 61.7 (22.8) | 0.26 |

| | | | | |
|---|---|---|---|---|
| Depression (Patient Health Questionnaire-9), mean (SD), points | 7.8 (6.5) | 8.7 (6.6) | 6.0 (5.7) | 0.43 |
| Stress (Perceived Stress Scale-14) , mean (SD), points | 26.0 (9.8) | 27.1 (9.9) | 23.6 (9.0) | 0.36 |

Abbreviations: EQ-5D, EuroQoL 5D; ESSI, ENRICHD Social Support Inventory; PCI, percutaneous coronary interventions; PHQ-9, Patient Health Questionnaire; PSS, Perceived Stress Scale; SAQ, Seattle Angina Questionnaire; SD, standardized difference; SF-12, Short Form-12.

Supplemental Table 5. Sex difference in AMI presentation and treatment

| | Total N=2,979 | Women N=2,007 | Men N=972 | Standardized Difference |
|---|---|---|---|---|
| Chest pain as primary symptom (%) | 2,600 (87.3) | 1,733 (86.3) | 867 (89.2) | 0.09 |
| Delayed hospital presentation (%) | 1,319 (44.5) | 951 (47.6) | 368 (38.0) | 0.19 |
| Killip class at arrival | | | | 0.12 |
| 1 (%) | 2,705 (90.8) | 1,810 (90.2) | 895 (92.1) | |
| 2 (%) | 83 (2.8) | 66 (3.3) | 17 (1.7) | |
| 3 (%) | 22 (0.7) | 18 (0.9) | 4 (0.4) | |
| 4 (%) | 13 (0.4) | 10 (0.5) | 3 (0.3) | |
| GRACE score, mean (SD), points | 74.0 (62.0-87.0) | 75.0 (63.0-88.0) | 73.5 (61.0-86.0) | 0.13 |
| Type of MI | | | | 0.24 |
| STEMI (%) | 1,483 (49.8) | 920 (45.8) | 563 (57.9) | |
| NSTEMI (%) | 1,496 (50.2) | 1,087 (54.2) | 409 (42.1) | |
| MINOCA (%) | 257 (9.6) | 232 (12.8) | 25 (2.9) | 0.34 |
| VIRGO Taxonomy | | | | 0.41 |
| 1 (%) | 2,318 (77.8) | 1,501 (74.8) | 817 (84.1) | |
| 2a (%) | 33 (1.1) | 26 (1.3) | 7 (0.7) | |
| 2b (%) | 54 (1.8) | 45 (2.2) | 9 (0.9) | |

| | | | | |
|---|---|---|---|---|
| 3a (%) | 82 (2.8) | 76 (3.8) | 6 (0.6) | |
| 3b (%) | 145 (4.9) | 128 (6.4) | 17 (1.7) | |
| 4 (%) | 30 (1.0) | 28 (1.4) | 2 (0.2) | |
| 5 (%) | 17 (0.6) | 15 (0.7) | 2 (0.2) | |
| Primary PCI for STEMI (%) | 1,135 (77.3) | 711 (78.0) | 424 (76.1) | 0.13 |
| Door to balloon, mean (SD), minutes | 116.8 (144.4) | 124.8 (153.6) | 103.9 (127.3) | 0.04 |
| Conservative therapy for AMI (%) | 89 (3.0) | 67 (3.3) | 22 (2.3) | 0.14 |
| Periprocedural complications | | | | 0.06 |
| MI re-infarction (%) | 28 (0.9) | 21 (1.1) | 7 (0.7) | 0.03 |
| Cardiac arrhythmias (%) | 205 (6.9) | 132 (6.6) | 73 (7.5) | 0.04 |
| Heart Failure (%) | 215 (7.3) | 160 (8.1) | 55 (5.7) | 0.09 |
| Bleeding (%) | 197 (6.6) | 134 (6.7) | 63 (6.5) | 0.01 |
| Length of stay, mean (SD), minutes | 4.2 (3.9) | 4.4 (4.2) | 3.9 (3.3) | 0.11 |
| Discharge medications (%) | | | | |
| Aspirin (%) | 2,782 (93.4) | 1,859 (92.6) | 923 (95.0) | 0.09 |
| Clopidogrel (%) | 2,052 (68.9) | 1,355 (67.5) | 697 (71.7) | 0.09 |
| Statins (%) | 2,739 (91.9) | 1,814 (90.4) | 925 (95.2) | 0.11 |
| Beta-blockers (%) | 2,713 (91.1) | 1,798 (89.6) | 915 (94.1) | 0.18 |
| ACEi / ARBs (%) | 1,915 (64.3) | 1,229 (61.2) | 686 (70.6) | 0.16 |
| CCBs (%) | 148 (5.0) | 122 (6.1) | 26 (2.7) | 0.20 |

Abbreviations: ACEi , angiotensin converting enzyme inhibitors; AMI, acute myocardial infarction; ARBs, angiotensin receptor blockers; CCBs, Calcium channel blockers; NSTEMI, non ST myocardial infarction; SD, standard deviation, STEMI, ST-elevated myocardial infarction

Supplemental Table 6. Timing of 1-year outcomes presented as days from discharge among young women and men

| | Total N=2,979 | Women N=2,007 | Men N=972 | Standardized Difference |
|---|---|---|---|---|
| All-cause deaths | 126 (47-273) | 126 (51-272) | 126 (17-326) | 0.11 |
| All-cause hospitalization | 70 (18-180) | 71.5 (20-188) | 68 (14-162) | 0.10 |
| MI-related | 83 (23-179) | 80.5 (24-189) | 85 (18-172) | 0.08 |
| Other cardiac/stroke | 49 (12-162) | 59 (15-179) | 29 (5-121) | 0.20 |
| Non-cardiac | 68 (16-191) | 65.5 (15-196) | 83.5 (27-167) | 0.05 |

Supplemental Table 7. Sex difference in invasive procedures performed during hospitalization

| | Total N=2,979 | Women N=2,007 | Men N=972 | Standardized Difference |
|---|---|---|---|---|
| Coronary catheterization | 283 (31.3%) | 211 (30.8%) | 72 (32.6%) | 0.04 |
| POBA | 41 (4.5%) | 27 (3.9%) | 14 (6.3%) | 0.11 |
| Stent placement | 123 (13.6%) | 95 (13.9%) | 28 (12.7%) | 0.04 |
| CABG | 26 (2.9%) | 18 (2.6%) | 8 (3.6%) | 0.06 |
| Pacemaker insertion | 2 (0.2%) | 1 (0.1%) | 1 (0.5%) | 0.07 |
| ICD placement | 9 (1.0%) | 5 (0.7%) | 4 (1.8%) | 0.11 |

Abbreviations: CABG: coronary artery bypass graft surgery, ICD: intracardiac defibrillator, POBA: plain old balloon angioplasty
