## Supplementary material for "Sex Difference in Causes and Timing of One-Year Outcomes Among Young Acute Myocardial Infarction Patients; Results from the VIRGO Study": VIRGO Participating Sites

| Site Name | City | State |
|---|---|---|
| Sentara Hospitals | Norfolk | VA |
| Yale-New Haven Hospital | New Haven | CT |
| Munson Medical Center | Traverse City | MI |
| Cardiovascular Research Foundation of Louisiana | Baton Rouge | LA |
| Mercy General Hospital | Sacramento | CA |
| Spectrum Health | Grand Rapids | MI |
| Washington University School of Medicine | Saint Louis | MO |
| Baylor Research Institute | Dallas | TX |
| St. John Hospital & Medical Center | Detroit | MI |
| Pepin Heart Hospital | Tampa | FL |
| Rhode Island Hospital | Providence | RI |
| St Mary's Medical Center | Huntington | WV |
| The Indiana Heart Hospital | Indianapolis | IN |
| St John's Mercy Medical Research Institute | Springfield | MO |
| St Francis Hospital and Medical Center | Hartford | CT |
| Parkview Hospital | Fort Wayne | IN |
| St. Elizabeth's Hospital | Belleville | IL |
| New Hanover Regional Medical Center | Wilmington | NC |
| Presbyterian Heart Group | Albuquerque | NM |
| The Ohio State University Medical Center | Columbus | OH |
| University of Southern California | Los Angeles | CA |
| Memorial Health System | Springfield | IL |
| Henry Ford Hospital | Detroit | MI |
| Washington Hospital Center | Washington, DC | |
| Memorial Medical Center | Modesto | CA |
| Montefiore / Albert Einstein Medical Center | Bronx | NY |
| Virginia Commonwealth University | Richmond | VA |
| Geisinger Clinic - Cardiology | Danville | PA |
| Fletcher Allen Health Care | Burlington | VT |

| | | |
|---|---|---|
| University of Texas Southwestern at Dallas (UTSW). | Dallas | TX |
| BryanLGH Heart Institute Research Services | Lincoln | NE |
| Bridgeport Hospital | Bridgeport | CT |
| Cardiovascular Clinical Research Center | New York | NY |
| Oakwood Hospital and Medical Center | Dearborn | MI |
| MAHI, St Luke's Hospital | Kansas City | MO |
| Long Beach Memorial Med Ctr | Long Beach | CA |
| Winthrop University Hospital | Mineola | NY |
| St. Rita's Medical Center | Lima | OH |
| Northshore University Health System | Evanston | IL |
| University of Michigan Health Systems | Ann Arbor | MI |
| Sutter Medical Center Sacramento | Sacramento | CA |
| Presbyterian Hospital - Mid Carolina Cardiology | Charlotte | NC |
| University of Alabama at Birmingham | Birmingham | AL |
| Stanford University Medical Center | Stanford | CA |
| University of Iowa Hospitals and Clinics | Iowa City | IA |
| Samaritan Heart and Vascular Institute | Corvallis | OR |
| Central Maine Heart and Vascular Institute | Lewiston | ME |
| Forsyth Medical Center | Winston-Salem | NC |
| Waukesha Memorial Hospital | Waukesha | WI |
| Torrance Memorial Medical Center | Torrance | CA |
| University of Chicago Medical Center | Chicago | IL |
| Hamot Medical Center | Erie | PA |
| Providence Healthcare Network | Waco | TX |
| Overlook Medical Center | Summit | NJ |
| Summa Health System (Akron City Hospital) | Akron | OH |
| St. Vincent Hospital & Health Services | Indianapolis | IN |
| D. Guthrie Foundation for Educ & Research | Sayre | PA |
| The International Heart Institute of Montana | Missoula | MT |
| Leonard J. Chabert Medical Center | Houma | LA |
| Boston University Medical Center | Boston | MA |
| Georgia Health Sciences University | Augusta | GA |

| | | |
|---|---|---|
| Good Samaritan Hospital | Kearney | NE |
| Saddleback Memorial Medical Center | Laguna Hills | CA |
| Cotton-O'Neil Clinical Research Center | Topeka | KS |
| St. Joseph's Medical Center | Stockton | CA |
| St Charles Health System | Bend | OR |
| SUNY Downstate Medical Center | Brooklyn | NY |
| The Valley Hospital | Ridgewood | NJ |
| Stony Brook University Medical Center | Stony Brook | NY |
| Minneapolis Heart Institute | Minneapolis | MN |
| Sharp Grossmont Hospital | La Mesa | CA |
| University of North Carolina at Chapel Hill | Chapel Hill | NC |
| Orlando Health | Orlando | FL |
| Butler Health System | Butler | PA |
| Krannert Heart Institute - Indiana University | Indianapolis | IN |
| Jefferson Regional Medical Center | Jefferson Hills | PA |
| St John's Regional Medical Center | Joplin | MO |
| Baptist Hospital of Miami (BCVI) | Miami | FL |
| Beth Israel Deaconess Medical Center | Boston | MA |
| Los Robles Hospital and Medical Center | Thousand Oaks | CA |
| Denver Health Medical Center | Denver | CO |
| Morehouse School of Medicine | Atlanta | GA |
| Allegheny General Hospital | Pittsburgh | PA |
| Mercy Hospital and Medical Center | Chicago | IL |
| Jamaica Hospital Medical Center | Jamaica | NY |
| Mount Clemens Regional Medical Center | Mount Clemens | MI |
| Citrus Memorial Health Systems | Inverness | FL |
| Providence Hospital | 221 Mobile | AL |
| Cedars-Sinai Medical Center | Los Angeles | CA |
| Genesis Medical Center | Davenport | IA |
| Parker Adventist Hospital | Denver | CO |
| California Medical Center | San Francisco | CA |
| Cox South Hospital | Springfield | MO |

| | | |
|---|---|---|
| Hackensack University Medical Center | Hackensack | NJ |
| Meriter Hospital | Madison | WI |
| Baptist Hospital, Inc.- Pensacola | Gulf Breeze | FL |
| Pinnacle Health Cardiovascular Institute (MHVG) | Wormleysburg | PA |
| Sparrow Hospital | Lansing | MI |
| Heartland Cardiovascular Consultants | St. Joseph | MO |
| Hospital of St. Raphael | New Haven | CT |
| University of Kansas Hospital | Kansas City | KS |
| Danbury Hospital | Danbury | CT |
| Glacier View Cardiology | Kalispell | MT |
| University of Minnesota | Minneapolis | MN |
